# Safety and microbiological activity of a combined phage cocktail in persons with cystic fibrosis colonized with *Achromobacter*: Study protocol for a phase 1, investigator-initiated, randomized clinical trial

**DOI:** 10.64898/2026.11.22.26368705

**Authors:** Justin Massey, Kailey Hughes-Kramer, Sarah Snook Barrett, Breuna Bishop, Scott Rothenberger, Saima Aslam, Fernanda Silveira, Joseph Pilewski, Keven Robinson, Anna Zemke, Ryan Shields, Madison Stellfox, Daria Van Tyne, Ghady Haidar

## Abstract

**Background:** *Achromobacter* are a genus of difficult-to-treat, frequently multi-drug-resistant Gram-negative bacteria that can cause severe infections in people with cystic fibrosis (pwCF). Bacteriophages are an understudied yet promising therapy for these and other antibiotic-resistant bacteria. However, data are largely limited to case reports or series, and clinical trials among pwCF to date have not evaluated *Achromobacter*.

**Methods:** This is an investigator-initiated, open-label, randomized phase 1 pilot trial to evaluate the safety, tolerability, and biological effects of AchromoPhage, a cocktail of four genetically distinct, obligately lytic *Achromobacter*-targeting phages (phiACH01, phiACH04, phiACH06, phiACH07) with broad activity against *Achromobacter* species, in 12 adult participants with CF and pulmonary *Achromobacter* infection. Twelve participants will be randomized into three groups to receive a total of three, increasing, weekly doses of the AchromoPhage cocktail via one of three routes: 1) intravenous, 2) via inhalation, or 3) or intravenously and inhaled. Participation in the trial will span approximately 7 weeks, which will consist of outpatient phage dose administrations, safety and symptom monitoring, and longitudinal collection of biospecimens for protocol-specified and exploratory analyses through Day 42. The primary safety endpoint is the occurrence of treatment-related Grade 4 or greater adverse events. Secondary endpoints include longitudinal changes in FEV1 by spirometry, longitudinal changes in health-related quality of life, pharmacokinetics of each of the four phages in blood and the respiratory tract, and emergence of a phage neutralizing antibody response in serum.

**Discussion:** While existing case anecdotes and observational studies suggest that phages are safe and may have a role in treating human infections, there is a paucity of clinical trials studying the adverse effect profile and basic pharmacological parameters of phages. This trial will study the safety and explore the pharmacokinetic and other properties of a four-phage cocktail in patients with CF and chronic *Achromobacter* lung infection.

**Trial registration:** ClinicalTrials.gov NCT0727590. Registered on 12/10/2025

**Structured summary:** 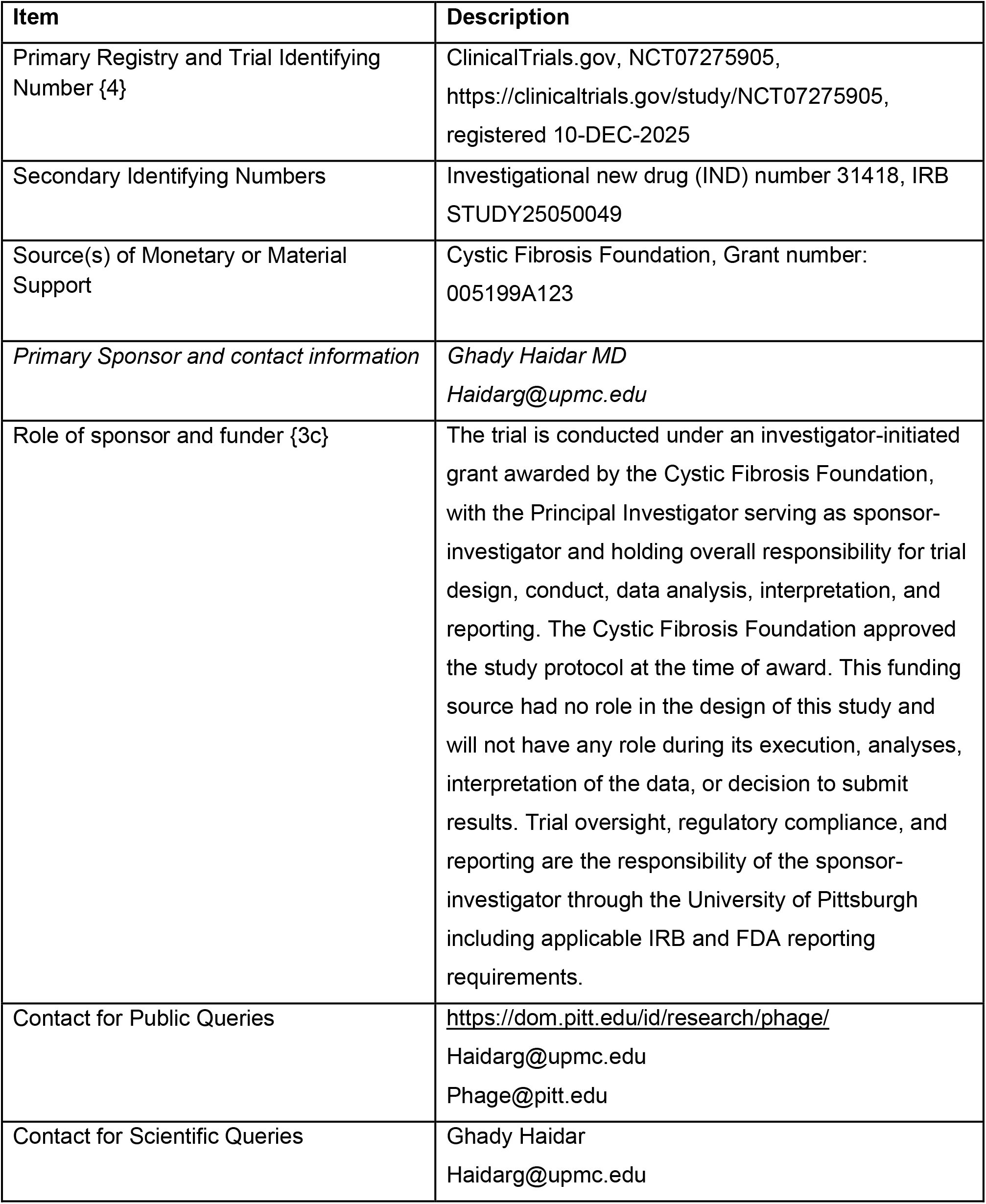

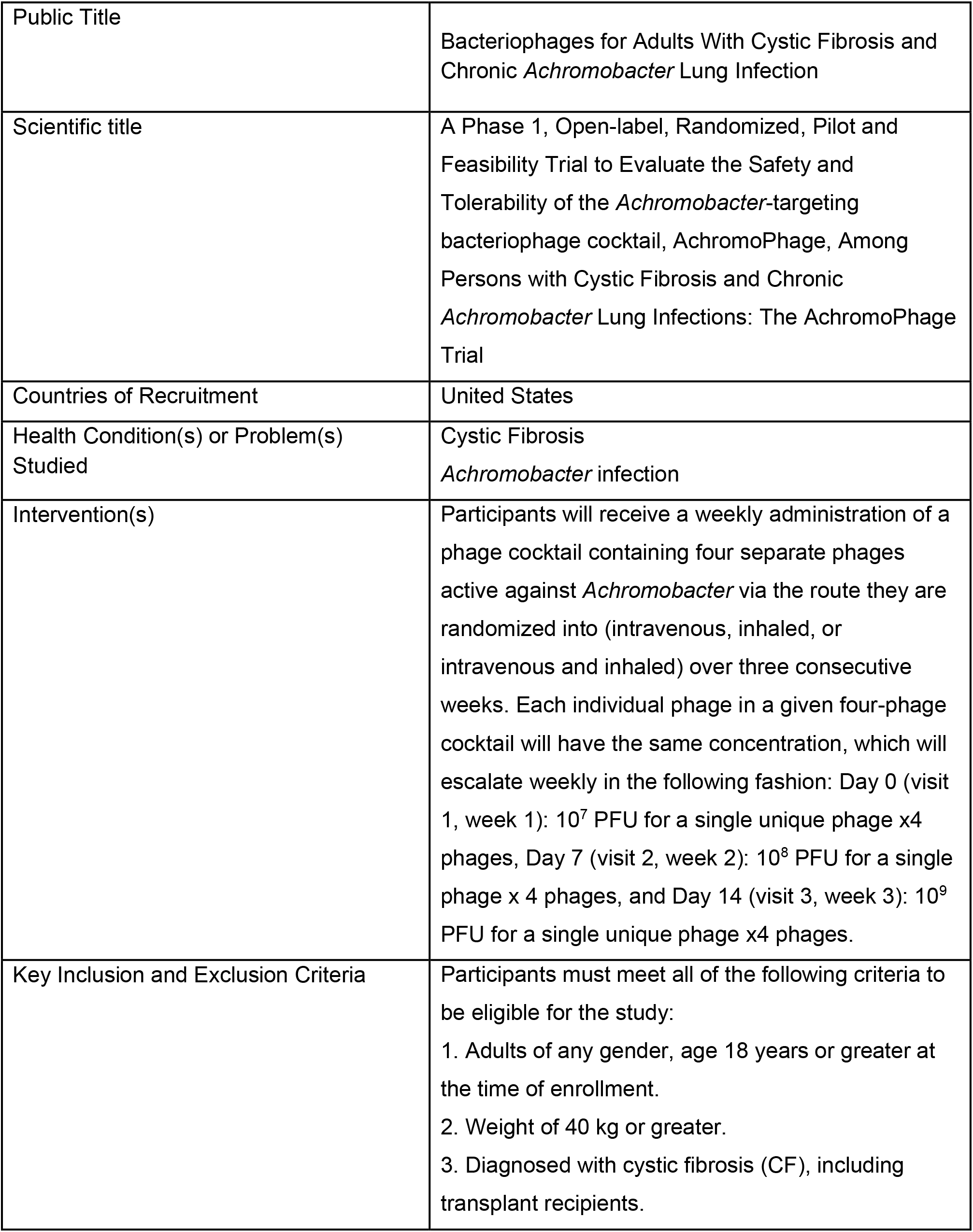

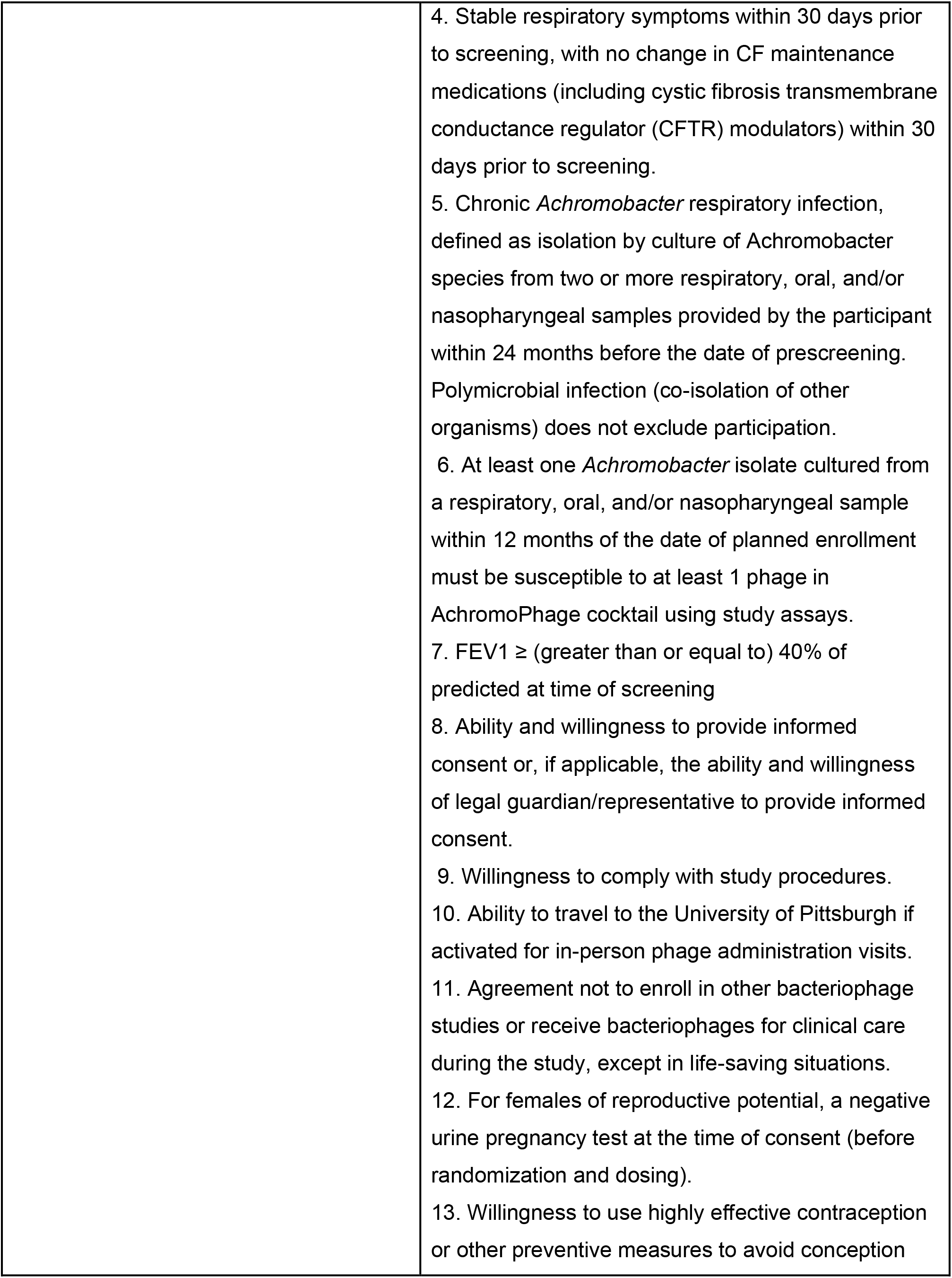

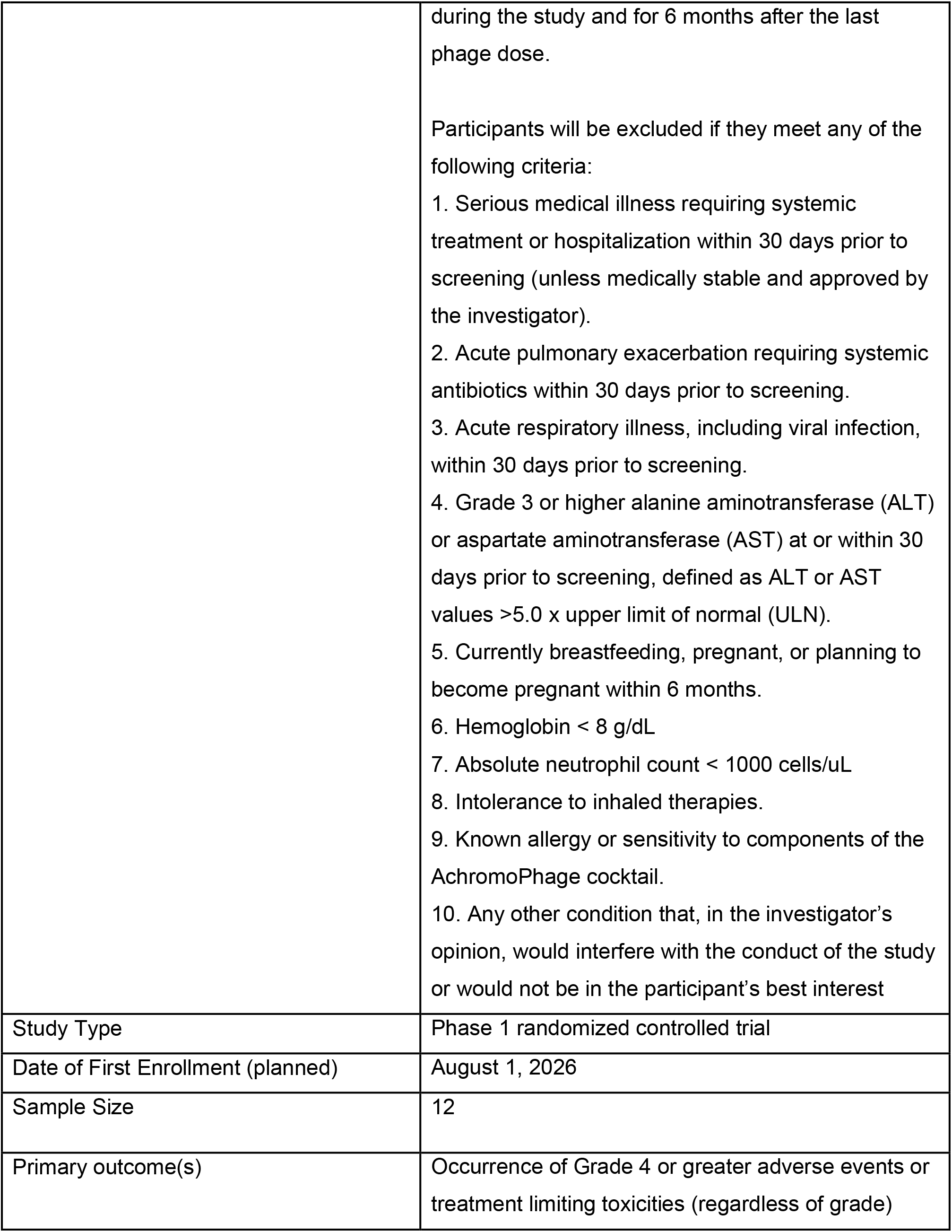

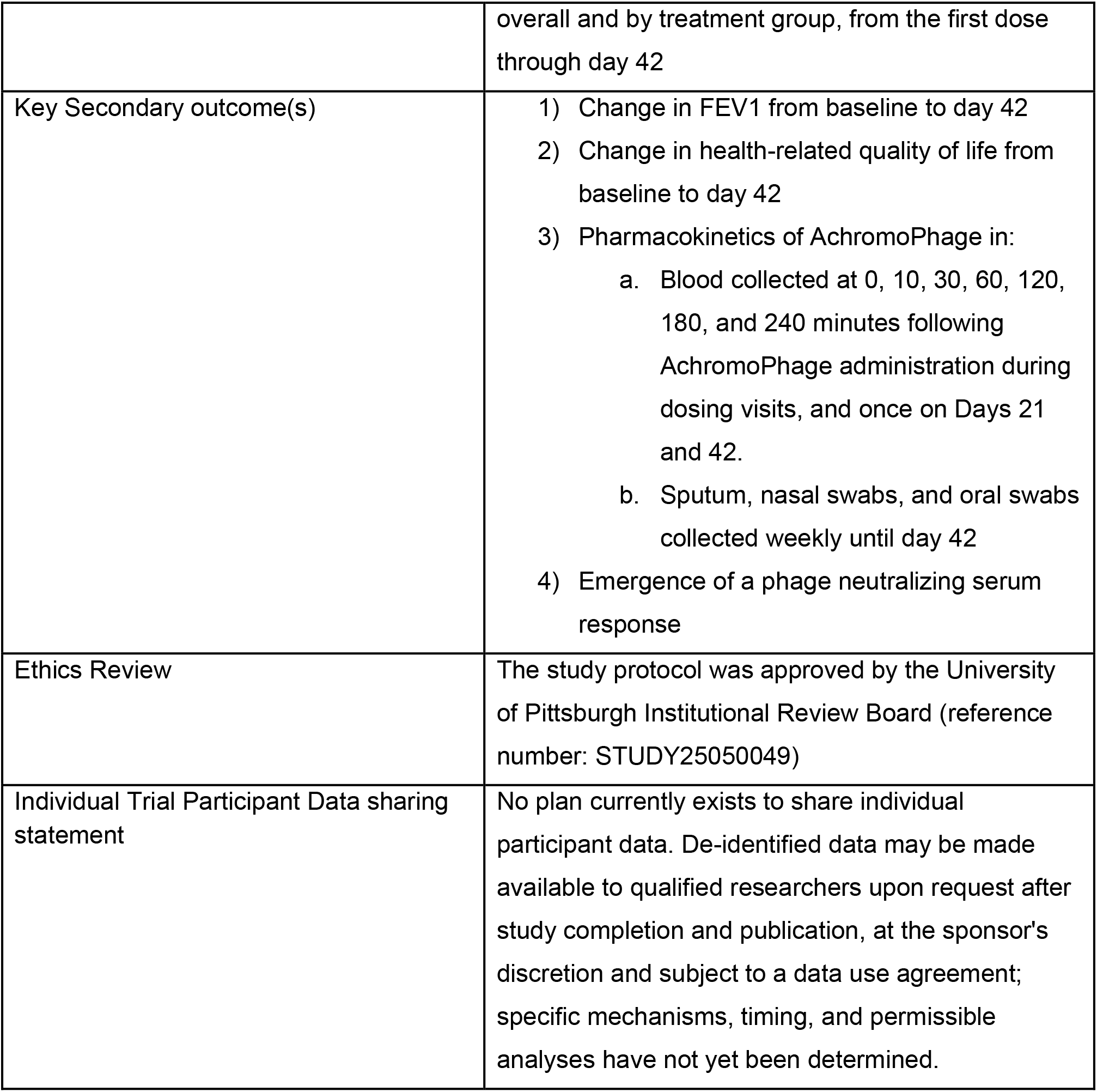

**Protocol version:** AchromoPhage Protocol August 26, 2026. Version 2.0

## Introduction

### Background and rationale

Despite the introduction of cystic fibrosis transmembrane conductance regulator (CFTR) modulator therapy(1–3), multidrug-resistant organisms (MDROs) continue to cause life-threatening infections in persons with cystic fibrosis (pwCF). Unlike *Pseudomonas aeruginosa* and *Burkholderia cepacia* complex infection, which are two of the most formidable and well-described pathogens in pwCF(3,4), the burden and consequences of *Achromobacter* spp. as only been recently defined. *Achromobacter* is estimated to affect 10–13% of pwCF and is associated with worsening forced expiratory volume in the 1st second (FEV1), accelerated FEV1 decline, and increased risk of pulmonary exacerbations, hospitalizations, and death (5–8). A growing proportion of *Achromobacter* isolates are multi- or pan-drug resistant (5,6,9), resulting in limited treatment options for patients. Unfortunately, *Achromobacter* has remained a neglected target for antimicrobial development.

Bacteriophages (phages)—viruses that target and kill bacteria—have emerged as a potential strategy for difficult to treat and multidrug-resistant organism (MDRO) infections, including in pwCF(10–22). First used to treat bacterial infections in the early 20th century(10,11), phages offer several advantages over antibiotics: they multiply at the site of infection ("self-dosing"), are highly specific to particular bacterial strains, and are thought to be "microbiome-sparing"(23). Nonetheless, phages were largely abandoned in the United States and Europe after penicillin ushered in the antibiotic era in the mid-1940s. More than 75 years later, however, antibiotic resistance continues to emerge unchecked. Even next-generation antimicrobuals developed as “last resort countermeasures” are met with resistance almost as quickly as they reach the clinic (24–29), a problem that is compounded by the absence of new antimicrobial classes. Accordingly, there has been renewed interest in the therapeutic potential of phage therapy, particularly in the United States.

Over the past decade, case reports and case series have suggested that phage therapy is safe and may provide clinical benefit in select individuals, including a small number of patients who were treated for *Achromobacter* (30–33). Nonetheless, much of the contemporary experience continues to be derived from single-patient, last-resort administrations (30,32). While informative and – importantly – sometimes beneficial to the patient, these experiences are not designed to generalize knowledge or advance the field. Furthermore, because phages have largely bypassed the traditional drug development pathway, fundamental parameters—pharmacokinetics, tissue distribution, immunogenicity, and other biological effects—remain poorly defined. Clinical trials are needed to develop safe, evidence-based protocols for phage therapy. A favourable regulatory and scientific landscape has enabled the launch of several early-phase phage trials. Among pwCF, several trials have studied nebulized or intravenous Pseudomonas-targeting phage for chronic *P. aeruginosa* infection, (including NCT05453578 and NCT04596319) with early results suggesting feasibility and an acceptable safety profile (34). Despite this progress, *Achromobacter* remains a neglected CF pathogen and is not the focus of industry-sponsored phage trials.

Since 2020, the Pittsburgh Phage Program (P3) (35), a team of academic investigators with clinical and scientific phage therapy expertise, has provided phage to more than 70 patients with difficult-to-treat infections through the FDA’s compassionate use pathway (36). This experience has directly informed our approach to key elements in the design of this trial, including phage dosing, route, duration, and assessment of pharmacokinetic, pharmacodynamic, and immunologic responses. The overarching goal of the AchromoPhage Trial is to determine the safety and tolerability of escalating-dose bacteriophage therapy, administered intravenously (IV) and by inhalation in individuals with chronic *Achromobacter* infection, and to generate the pharmacokinetic, pharmacodynamic, and immunologic data needed to inform the design of a future phase 2 trial.

### Explanation for the choice of comparator

This trial has no placebo or standard-of-care comparator; participants are randomized among three routes of AchromoPhage administration (inhaled, intravenous, or both), and comparisons between arms are descriptive. A placebo arm was not included because *Achromobacter* infects only 10–13% of pwCF, compared with over 60% for *Pseudomonas aeruginosa*, and adding one would have required a larger sample to ensure that 12 participants receive active product. As the primary objective of this first-in-human trial is safety and tolerability, an uncontrolled design was considered appropriate. All participants continue their usual CF care, including maintenance inhaled antimicrobials, throughout the trial.

These three treatment arms (N = 4 participants each) were selected as the optimal route for delivering phage to an infected lung is unknown (37). Data evaluating different methods of phage delivery in respiratory infections are sparse(38). Studies in other respiratory diseases have demonstrated that inhaled medications are effective, such as inhaled nitric oxide and nebulized antibiotics for prevention of CF exacerbations (39,40). Inhaled phage would similarly allow for direct delivery of the drug product to the site of infection, but the foundational data to establish its safety and efficacy has not been systematically established. While IV phage achieves systemic delivery, it is unclear whether chronic ventilation/perfusion (V/Q) mismatching limits penetration of phage into poorly perfused lung regions. Systemic exposure may also increase the likelihood of an immune response to subsequent doses via a neutralizing antibody response(41). Compassionate use practices vary, but have often defaulted to combining IV and inhaled phage(42,43), reflecting empiricism rather than evidence of superiority over either route alone. Importantly, this trial is not designed to resolve which route is optimal; comparative safety and pharmacokinetic/pharmacodynamic data across arms will instead inform route selection and sample size for subsequent phase 2 trials.

### Objectives

Primary objective:

1. To assess the safety and tolerability of three escalating doses of AchromoPhage in adult pwCF with chronic *Achromobacter* infection across three different routes of administrations. *<u>Timepoints</u>: from baseline to day 42*.

Secondary objectives:

1. To evaluate changes in FEV1 (spirometry) before and after AchromoPhage administration. *<u>Timepoints</u>: from baseline to day 42*
2. To assess changes in health-related quality of life via the Cystic Fibrosis Questionnaire-Revised before and after AchromoPhage administration. *<u>Timepoints</u>: from baseline to day 42*
3. To characterize the pharmacokinetics of AchromoPhage in blood, sputum, nasal swabs, and oral swabs.

*a) Blood: Collected at 0-, 10-, 30-, 60-, 120-, 180-, and 240-minute intervals following AchromoPhage administration during dosing visits (Days 0, 7, and 14) and once on Days 21 and 42;*
*b) Sputum, nasal swabs, and oral swabs: Collected weekly (Days 0, 7, 14, 21, 28, 35, and 42).*
4. To assess the humoral immune response to the AchromoPhage cocktail. *<u>Timepoints: baseline and days 7, 14, 21, 28, and 42.</u>*

Exploratory objectives:

1. To evaluate the pharmacodynamics of AchromoPhage, focusing on changes in *Achromobacter* abundance, total bacterial abundance, and other phage-host-microbe interactions and dynamics
2. To examine the evolution and phage-pathogen susceptibility of *Achromobacter* and phages over time

### Description of trial estimand(s)

The primary estimand is defined below. Given the pilot nature of the trial, secondary and exploratory outcomes are summarized descriptively and no formal estimands are specified for them.

| Estimand Value | Description |
|---|---|
| Population | Adults with cystic fibrosis meeting the eligibility criteria. Broadly, individuals older than 18 years of age with stable respiratory symptoms and chronic *Achromobacter* infection that is susceptible to at least one phage within the AchromoPhage cocktail. |
| Treatment conditions | Three weekly AchromoPhage administrations with dose escalation (4×10<sup>7</sup>, 4×10<sup>8</sup>, and 4×10<sup>9</sup> PFU), delivered by one of three routes: inhaled alone, intravenous alone, or inhaled followed 60 minutes later by intravenous in the same visit. All participants continue usual CF care. |
| Variables of interest | - Occurrence in an individual participant of either a treatment-related Grade 4 or higher adverse event or a treatment-limiting toxicity of any grade, from the first phage dose through Day 42 (binary composite). |
| Handling of intercurrent events | - *Discontinuation of phage dosing before all three doses* (for a stopping rule, intolerance, pregnancy, or participant request): treatment policy strategy, with all events occurring through Day 42 counted regardless of the number of doses received; participants who do not complete all three doses and the Visit 4 assessments are replaced.<br>- *Acute pulmonary exacerbation and initiation of new systemic or inhaled antibiotics*: treatment policy strategy; exacerbations are also recorded as adverse events of special interest.<br>- *Rescue bronchodilator use*: treatment policy strategy, with use documented.<br>- *Withdrawal or loss to follow-up*: treated as missing data under a missing-at-random assumption. |
| Population level summary of the comparison between treatment conditions | Incidence of the composite endpoint, estimated for all participants combined and separately by route, with one-sided 90% confidence intervals. Between-route differences are estimated as differences in least-squares means with two-sided 90% confidence intervals, without adjustment for multiplicity; no formal hypothesis testing is performed. |

## Methods: Patient and public involvement, and trial design

### Patient and public involvement

Patients were involved in the review of this trial through the Cystic Fibrosis Foundation’s grant review process, which includes community reviewers who are people with cystic fibrosis. Their review informed the funded design of the trial. The Foundation also reviewed and agreed the participant compensation plan. Patients and members of the public were not otherwise involved in the conduct of the trial, although the requirement for in-person attendance at the two non-dosing follow-up visits was relaxed during enrollment after out-of-state participants found the additional travel burdensome. Trial results will be disseminated to the CF community in addition to publication in a peer-reviewed journal.

### Trial design

This is a phase 1 randomized control trial of three increasing weekly doses of a 4-phage cocktail (AchromoPhage) by either 1) IV, 2) inhaled, or 3) IV and inhaled route in 12 clinically stable adult participants with cystic fibrosis who have chronic airway colonization with *Achromobacter*. The primary study outcomes include the safety and tolerability of the 3 escalating doses of AchromoPhage through the initial dose into the final day of the trial (day 42).

## Methods: Participants, interventions and outcomes

### Trial setting

Persons with CF who meet eligibility criteria will be recruited from across the United States. Phage dosing visits must occur on site at UPMC in Pittsburgh, PA. The trial reimburses travel costs.

## Characteristics of the people who are needed for the trial

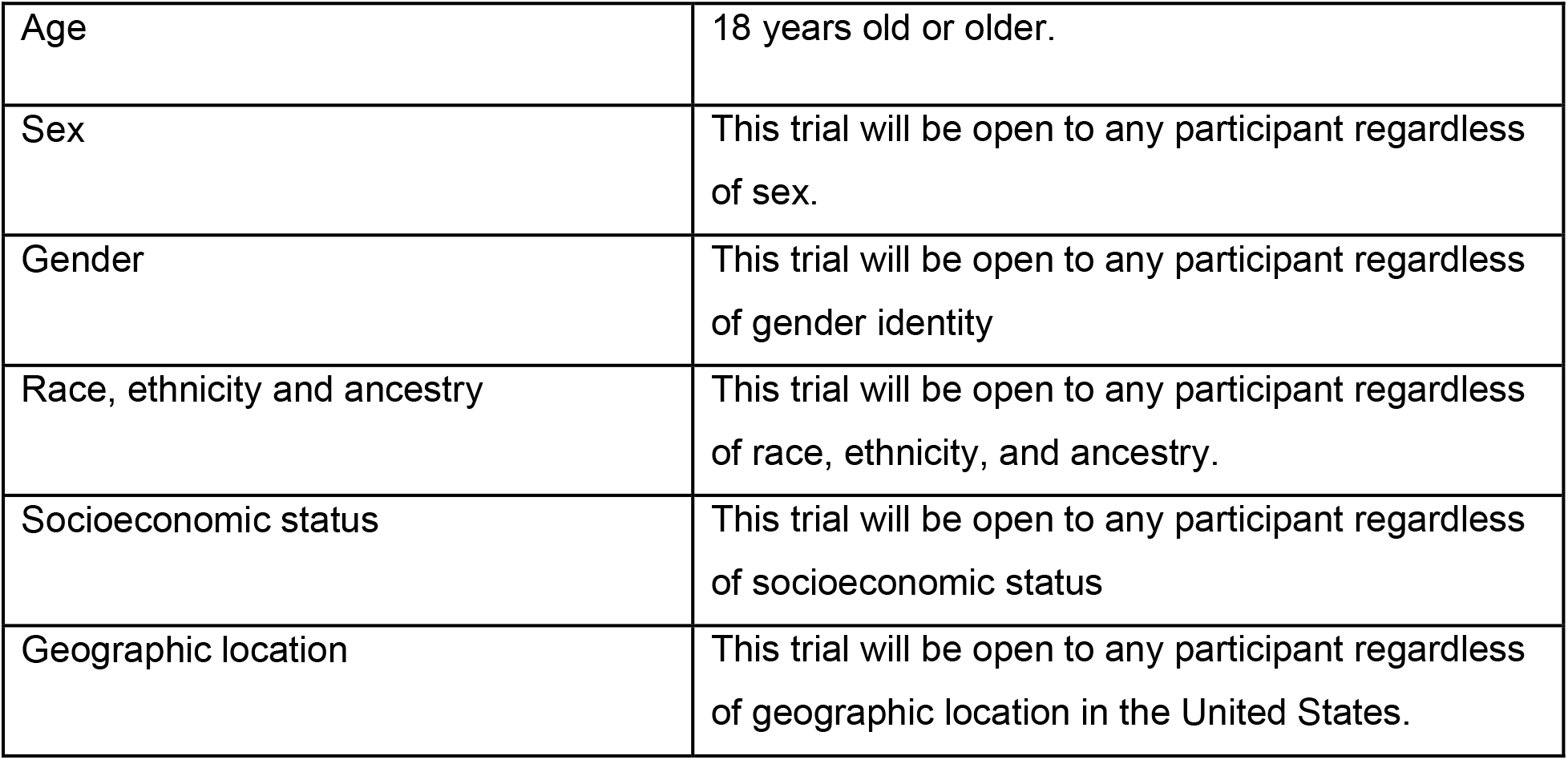
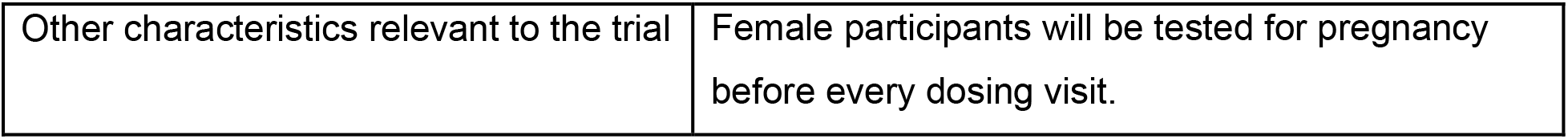

| Age | 18 years old or older. |
|---|---|
| Sex | This trial will be open to any participant regardless of sex. |
| Gender | This trial will be open to any participant regardless of gender identity |
| Race, ethnicity and ancestry | This trial will be open to any participant regardless of race, ethnicity, and ancestry. |
| Socioeconomic status | This trial will be open to any participant regardless of socioeconomic status |
| Geographic location | This trial will be open to any participant regardless of geographic location in the United States. |
| Other characteristics relevant to the trial | Female participants will be tested for pregnancy before every dosing visit. |

## Eligibility criteria for participants

Participants must meet all of the following inclusion criteria to be eligible for the study:

1. Adults of any gender, age 18 years or greater at the time of enrollment.
2. Weight of 40 kg or greater.
3. Diagnosed with cystic fibrosis, including transplant recipients.
4. Stable respiratory symptoms within 30 days prior to screening, with no change in CF maintenance medications (including cystic fibrosis transmembrane conductance regulator (CFTR) modulators) within 30 days prior to screening.
5. Chronic *Achromobacter* respiratory infection, defined as isolation by culture of *Achromobacter* species from two or more respiratory, oral, and/or nasopharyngeal samples provided by the participant within 24 months before the date of prescreening. Polymicrobial infection (co-isolation of other organisms) does not exclude participation.
6. At least one *Achromobacter* isolate cultured from a respiratory, oral, and/or nasopharyngeal sample within 12 months of the date of planned enrollment must be susceptible to at least 1 phage in AchromoPhage cocktail using study assays.
7. FEV1 ≥ (greater than or equal to) 40% of predicted at time of screening
8. Ability and willingness to provide informed consent or, if applicable, the ability and willingness of legal guardian/representative to provide informed consent.
9. Willingness to comply with study procedures.
10. Ability to travel to the University of Pittsburgh if activated for in-person phage administration visits.
11. Agreement not to enroll in other bacteriophage studies or receive bacteriophages for clinical care during the study, except in life-saving situations.

Participants who meet any of the following exclusion criteria may not be enrolled in the study:

1. Serious medical illness requiring systemic treatment or hospitalization within 30 days prior to screening (unless medically stable and approved by the investigator).
2. Acute pulmonary exacerbation requiring systemic antibiotics within 30 days prior to screening.
3. Acute respiratory illness, including viral infection, within 30 days prior to screening.
4. Grade 3 or higher alanine aminotransferase (ALT) or aspartate aminotransferase (AST) at or within 30 days prior to screening, defined as ALT or AST values >5.0x upper limit of normal (ULN).
5. Currently breastfeeding, pregnant, or planning to become pregnant within 6 months.
6. Hemoglobin < 8 g/dL
7. Absolute neutrophil count < 1000 cells/uL
8. Intolerance to inhaled therapies.
9. Known allergy or sensitivity to components of the AchromoPhage cocktail.
10. Any other condition that, in the investigator’s opinion, would interfere with the conduct of the study or would not be in the participant’s best interest

## Eligibility criteria for sites and those delivering interventions

Participants are recruited nationally from CF centers across the United States, but all phage administration visits take place on site at the University of Pittsburgh Medical Center (UPMC) in Pittsburgh, Pennsylvania, with travel and lodging covered by the trial. The University of California San Diego may be activated as a second enrolling site only if accrual is slower than anticipated. No formal site-level eligibility criteria are applied. Sites must have an investigational drug pharmacy able to store, prepare, and dispense AchromoPhage under the specified conditions and to perform randomization through REDCap, a vibrating mesh nebulizer for inhaled dosing, capacity for monitored dosing visits of approximately four hours, spirometry meeting American Thoracic Society and European Respiratory Society standards, and access to a clinical microbiology laboratory for retrieval of remnant specimens and isolates.

All study personnel complete human subjects protection and ICH Good Clinical Practice training. Phage doses are prepared by investigational drug pharmacists and administered by qualified medical personnel; sputum induction, where performed, is conducted by a respiratory therapist or other qualified personnel; and indwelling catheters are accessed only by credentialed staff.

## Who will take informed consent?

Written informed consent will be obtained from all participants before any study-specific procedure is performed. The institutional review board determines which study personnel are authorized to conduct the consent discussion and obtain signatures, in accordance with FDA requirements for licensed healthcare professional involvement, state and local requirements, and applicable law. Consent may be obtained either in person or by remote contact using IRB-approved consent forms and discussion aids; electronic consent is obtained using 21 CFR Part 11–compliant platforms. Consent forms are provided to participants in advance where possible, are written at a reading level no higher than eighth grade, and translations and an interpreter are available as needed. Participants receive a copy of the signed form, and consent, randomization, and the first phage dose all occur on Day 0. To confirm the phage susceptibility eligibility criterion, limited consent for biospecimen collection may be obtained remotely, including by telephone. Where a potential participant is unable to consent on their own behalf, sites follow their standard procedures for obtaining consent from a legally authorized representative. Participants are re-consented if a protocol amendment affects the consent form, if new information emerges that could affect willingness to continue, or if otherwise required by the IRB.

## Additional consent provisions for collection and use of participant data and biological specimens

Biological specimens collected under this protocol will undergo testing as outlined in the study protocol (phage pharmacokinetics in blood and respiratory specimens, phage-neutralizing antibody responses, the microbiological impact of phage therapy in the airways, and additional research assays). Excess specimens will be stored indefinitely at the University of Pittsburgh in de-identified or coded form and may undergo future testing as outlined in the protocol. Any future research will comply with applicable regulatory requirements including informed consent, IRB oversight, and privacy protections in accordance with FDA regulations and applicable human subjects protection laws. Participants are separately offered optional consent for future genetic testing and for an optional longitudinal biospecimen component, under which remnant clinical Achromobacter isolates may be collected from before the study and for up to 24 months after the trial ends. Both are opt-in, and declining either does not affect participation in the main trial.

## Intervention and comparator

### Intervention and comparator description

AchromoPhage is a cocktail of four separate phages (phiACH01, phiACH04, phiACH06, and phiACH07) in equal concentrations. The cocktail is prepared in 1x phosphate buffered saline (PBS) containing 10mM MgSO4 and is tested to confirm the presence of expected phage DNA, as well as confirm minimal endotoxin content and ensure sterility.

Participants receive one administration per week for three consecutive weeks, with weekly dose escalation. Each phage is present at 1×10⁷, 1×10⁸, and 1×10⁹ PFU/mL at Visits 1, 2, and 3 respectively, giving total doses of 4×10⁷, 4×10⁸, and 4×10⁹ PFU per administration in the single-route arms. For the intravenous dose, the total 1mL vial will be drawn up and given via IV push. For the inhaled doses, the same 1mL vials will be mixed with 2mL of normal saline to prepare a total 3mL dose for inhalation by a vibrating mesh nebulizer. The vibrating mesh nebulizer was chosen over a jet nebulizer because it better preserves phage viability during nebulization (44). Participants in the combination arm will receive an inhaled dose, followed by an IV dose one hour after the inhaled dose in the same visit, so that the total dose per administration is twice that of the single-route arms. All products are stored at 2 to 8 degrees Celsius and protected from light, with a beyond-use date of 72 hours after compounding. Each dose will be verified to contain less than 5 endotoxin units per kilogram of body weight per hour, per study participant. There is no placebo or active comparator; the three arms differ only in route of administration. Detailed preparation and administration procedures are provided in the pharmacy manual and manual of procedures.

### Criteria for discontinuing or modifying allocated intervention/comparator

AchromoPhage doses are fixed for each visit, and no dose reductions are planned; dosing may only be withheld, delayed, or permanently discontinued. Adverse events (AEs) are graded using the Common Terminology Criteria for Adverse Events version 5.0. Discontinuation of AchromoPhage will generally occur for any of the following:

- Any treatment-related AE of grade 3 or higher
- Any treatment-related serious adverse event
- Any treatment-related AE of special interest (acute CF pulmonary exacerbation, severe allergic or anaphylactic reaction, unexpected organ-specific toxicity, or severe paradoxical reaction)
- A relative decline of 20% or more in percent predicted FEV1 after a dose, compared with the pre-dose value at that visit
- Phage intolerance, including grade 2 or higher bronchospasm, a fall in systolic blood pressure of 40 mmHg or more from the pre-dose value, symptomatic systolic blood pressure below 80 mmHg, symptomatic oxygen desaturation (SpO2 below 88% or a fall of more than 5% from baseline), or grade 2 cytokine release syndrome
- Any persistent or recurrent treatment-related AE that the investigator judges incompatible with continued dosing
- Pregnancy before completion of all three phage administrations

If pre-dose percent predicted FEV1 at a dosing visit has declined by 20% or more relative to the Visit 1 baseline, dosing will be withheld until the investigator completes an assessment. A body weight below 40 kg at a dosing visit will prompt clinical assessment but will not automatically require discontinuation.

These criteria are guidance. The final decision rests with the investigator, who may continue dosing after a documented risk-benefit assessment or if an alternative cause is identified. The exception is treatment-related myocarditis of grade 2 or higher, which will lead to immediate discontinuation. The site investigator will notify the Sponsor-Investigator within 24 hours of any event that meets a stopping criterion. Participants may also stop the intervention at their own request.

Participants who discontinue the intervention after at least one phage dose will be offered the opportunity to complete all remaining study procedures without further dosing. Randomization will continue until four participants in each arm have received all three phage doses and completed the Visit 4 assessments, so participants who do not reach this point will be replaced.

## Strategies to improve adherence to intervention/comparator

The study is designed to minimize participant burden in order to support recruitment, retention, and adherence to the trial. While phage administration must occur at UPMC, initial contact including pre-screening between potential participants and the study team may occur remotely. Initial screening sputum specimens can be mailed to the lab for phage susceptibility testing. Visits 4 and 7 may be conducted either in person or remotely, with a teleconference with the principal investigator if remote, and Visits 5 and 6 are remote; specimens and spirometry for remote visits are collected at home. Participants are reimbursed for travel to the study site from anywhere in the United States, including mileage, parking, airfare, and lodging as needed, and are compensated for each visit completed (up to $900 in total).

Every phage dose is administered on site by study personnel, so adherence to the intervention is directly observed and recorded in source documents and the electronic case report form (date, time, dose, route, and administering staff). Adherence to home procedures (spirometry, symptom diaries, the CFQ-R, specimen collection, and temperature monitoring) is supported by check-in calls the day after each dose and by visit reminders, and study staff review participant-completed documentation for completeness. Participants are considered to have received the intervention as planned if they receive all three phage doses and complete the Visit 4 assessments.

## Concomitant care permitted or prohibited during the trial

There is no restriction on concomitant care during the trial. Participants will be asked to perform their usual morning airway clearance.

## Outcomes

Our primary outcomes are a composite binary endpoint, encompassing the development of:

1. Any treatment-related grade 4 or greater AE
2. Any treatment-limiting AE regardless of grade.

Secondary outcomes evaluated in this study include:

1. Change in FEV1 from baseline compared to day 42
2. Changed in health-related quality of life from baseline to day 42
3. Pharmacokinetic analysis of AchromoPhage in:

a. Blood collected at 0, 10, 30, 60, 120, 180, and 240 minutes after phage administration on days 0, 7, and 14, as well as once on days 21 and 42. Participants in the combination arm will have two t=0 timepoints (one timepoint for the inhaled dose, and another before the intravenous dose)
b. Sputum, nasopharyngeal swabs, and oropharyngeal swabs collected weekly on days 0,7,14,21,28,35, and 42. Samples will be taken twice (pre and post phage administration) on days phage is administered.
4. Emergence of a phage neutralizing antibody response in the serum on days 0, 7,14,21,28, and 42

Exploratory outcomes in this trial will consist of:

1. Analysis of the change in *Achromobacter* abundance and total bacterial abundance, and other changes in the pulmonary microbiota profiles over time
2. Changes in *Achromobacter* susceptibility to antibiotics and phages over time

## Harms

Harms are assessed from informed consent through Visit 7, four weeks after the final phage dose. The trial is open label, so participants, care providers, and outcome assessors are aware of the assigned route.

Solicited events are assessed systematically at each study visit and at scheduled remote contacts after dosing visits. Participants are asked directly about fever above 38 °C, chills, lightheadedness, night sweats, dyspnea, chest tightness, wheezing or bronchospasm, increased oxygen requirement, cough, productive cough, hemoptysis, malaise or fatigue, anorexia or weight loss, sinus pain or discharge, laryngeal edema, pulmonary exacerbation, and a relative decline of 20% or more in percent predicted FEV1. Participants also complete symptom diaries and record home temperatures around dosing visits.

Systematic monitoring additionally includes serial vital signs and pulse oximetry during the post-dose observation period, spirometry before and after each dose, weekly complete blood count and comprehensive metabolic panel, and a baseline electrocardiogram. Acute CF pulmonary exacerbation (identified using the modified Fuchs criteria, with investigator adjudication), severe allergic or anaphylactic reaction, unexpected organ-specific toxicity, and severe paradoxical reaction are designated adverse events of special interest and require rapid communication to the Sponsor-Investigator.

Non-solicited events are captured from unprompted participant reports and review of the electronic medical record; events captured as solicited are not recorded again as unsolicited. All adverse events are graded using the National Cancer Institute’s Common Terminology Criteria for Adverse Events version 5.0 by the investigator or a qualified clinician, who also assesses relatedness to AchromoPhage; seriousness is determined according to 21 CFR 312.32 and expectedness by the Sponsor-Investigator. In this population, certain disease-related events are expected from the natural course of cystic fibrosis and may overlap with adverse events; these are tracked separately and reclassified as adverse events if they exceed their expected frequency, severity, or duration.

Serious adverse events are reported to the Sponsor-Investigator within 24 hours, with a detailed follow-up report within five calendar days; non-serious events are reported monthly and aggregated quarterly for the Data Safety Monitoring Board. Serious and unexpected suspected adverse reactions are reported to the FDA within seven calendar days if fatal or life-threatening and within 15 days otherwise. Harms unrelated to the study intervention include breach of confidentiality, fatigue from spirometry, and bruising or syncope from phlebotomy.

## Participant timeline

When identified, all potential study participants will be educated on the study including the purpose, risk, and aims of the trial. Eligibility criteria will be reviewed, and if eligible, their *Achromobacter* isolate will be screened for phage susceptibility under limited consent. If their isolate is susceptible to at least one of the phages in the AchromoPhage cocktail, the patient may provide full written consent and formally enroll in the trial. The participant will then be given additional information on the logistics of the trial and be randomized into one of three treatment arms on Day 0. The prescreening process may all be done remotely. Across all treatment arms, study participants will undergo phage treatment on the first three visits (Days 0, 7, and 14). Attendance to the study site is required for all three of these visits.

Research evaluations including blood, sputum, NP/OP swabs, and the Cystic Fibrosis Questionnaire-Revised (CFQR) will be taken during these visits and continue throughout follow-up. Following the phage administration visits, participants will be followed weekly for a total of seven weeks (four additional weeks/visits – V4-V7). Phages will not be administered during these follow up visits, and participants may choose to conduct these visits remotely. Sputum and swabs collected at home, along with the symptom diary and CFQ-R will be sent by the participant to the study investigators; research blood is collected remotely at visits 4 and 7. The schedule of enrollment, interventions, and assessments are shown in **Figure 1** and **Table 1**.

**Figure 1:**
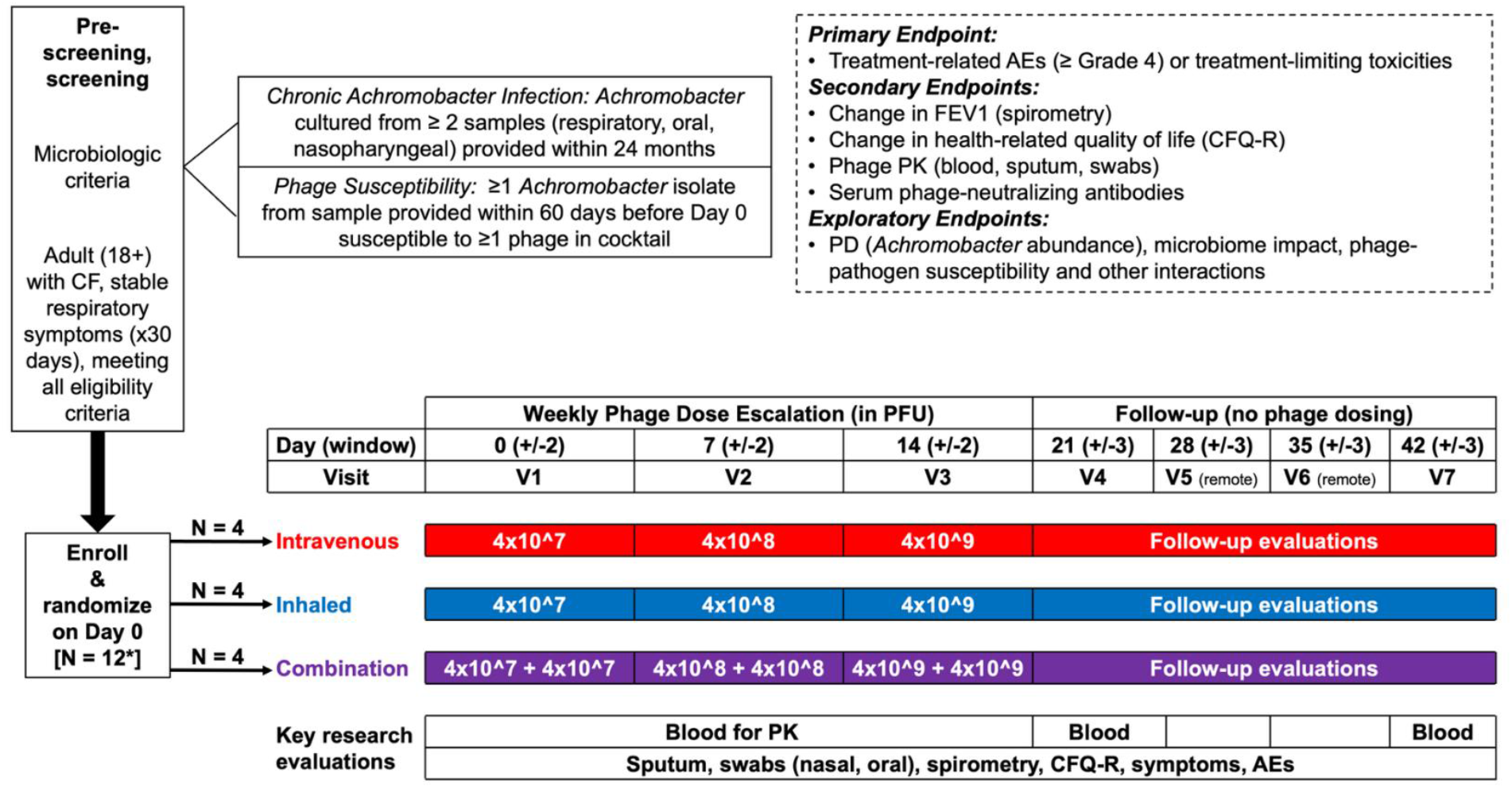
Schematic for participant timeline in the AchromoPhage trial. Following prescreening and confirmation of phage susceptibility, participants are enrolled and randomized into one of three treatment arms (IV alone, inhaled alone, and combination inhaled and IV arms). Participants in all three arms will have weekly, escalating doses of phage administered to them in their respective treatment arm. Following the weekly phage doses, participants will be monitored for four more weeks, with a total study duration of seven weeks.

**Table 1:**
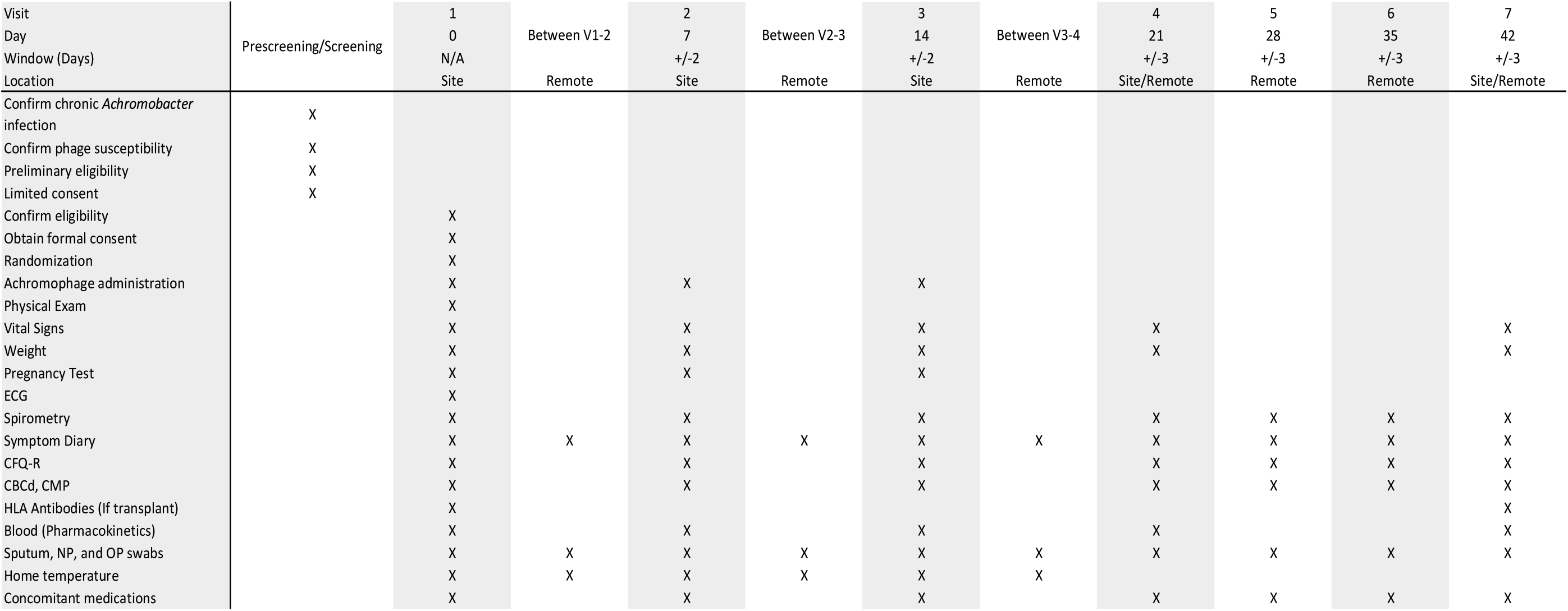
Schedule of events during each visit in the AchromoPhage trial. -Vitals signs are monitored throughout the entire phage dosing visit and once during non-dosing visits. -A pregnancy test will be taken for all females of reproductive age prior to all dosing visits. -Spirometry is performed before and after phage dosing on visits 1, 2, and 3, while only once on visits 4, 5, 6, and 7. -Participants in the combination arm will received a dose of inhaled phage first, then an IV dose 60 minutes after. -Blood for PK measurements will be taken pre-phage, then +10, +30, +60, +120, +180, and +240 minutes after phage administration in the IV and inhaled single route arms. In the combination arm, blood for PK will be taken pre-inhaled phage, then +10, +30, +60, +120, +180, +240, and +300 minutes after the IV phage is given

| Visit | Prescreening/Screening | 1 | Between V1-2 | 2 | Between V2-3 | 3 | Between V3-4 | 4 | 5 | 6 | 7 |
|---|---|---|---|---|---|---|---|---|---|---|---|
| Day |  | 0 |  | 7 |  | 14 |  | 21 | 28 | 35 | 42 |
| Window (Days) |  | N/A |  | +/-2 |  | +/-2 |  | +/-3 | +/-3 | +/-3 | +/-3 |
| Location |  | Site | Remote | Site | Remote | Site | Remote | Site/Remote | Remote | Remote | Site/Remote |
| Confirm chronic *Achromobacter* infection | X |  |  |  |  |  |  |  |  |  |  |
| Confirm phage susceptibility | X |  |  |  |  |  |  |  |  |  |  |
| Preliminary eligibility | X |  |  |  |  |  |  |  |  |  |  |
| Limited consent | X |  |  |  |  |  |  |  |  |  |  |
| Confirm eligibility |  | X |  |  |  |  |  |  |  |  |  |
| Obtain formal consent |  | X |  |  |  |  |  |  |  |  |  |
| Randomization |  | X |  |  |  |  |  |  |  |  |  |
| Achromophage administration |  | X |  | X |  | X |  |  |  |  |  |
| Physical Exam |  | X |  |  |  |  |  |  |  |  |  |
| Vital Signs |  | X |  | X |  | X |  | X |  |  | X |
| Weight |  | X |  | X |  | X |  | X |  |  | X |
| Pregnancy Test |  | X |  | X |  | X |  |  |  |  |  |
| ECG |  | X |  |  |  |  |  |  |  |  |  |
| Spirometry |  | X |  | X |  | X |  | X | X | X | X |
| Symptom Diary |  | X | X | X | X | X | X | X | X | X | X |
| CFQ-R |  | X |  | X |  | X |  | X | X | X | X |
| CBCd, CMP |  | X |  | X |  | X |  | X | X | X | X |
| HLA Antibodies (If transplant) |  | X |  |  |  |  |  |  |  |  | X |
| Blood (Pharmacokinetics) |  | X |  | X |  | X |  | X |  |  | X |
| Sputum, NP, and OP swabs |  | X | X | X | X | X | X | X | X | X | X |
| Home temperature |  | X | X | X | X | X | X |  |  |  |  |
| Concomitant medications |  | X |  | X |  | X |  | X | X | X | X |

## Sample size

The sample size of 12 is based on feasibility considerations, as is appropriate for pilot trials; no formal power calculation was performed. A sample size of 12 is expected to be sufficient to address tolerability/safety, given that our compassionate-use experience and the published literature both suggest that phage therapy is safe and well tolerated. Furthermore, in a Phase 1b/2 trial evaluating the safety and microbiological activity of bacteriophage therapy in persons with CF who are colonized with *Pseudomonas aeruginosa*(34), the Phase 1 arm of the trial assigned two eligible subjects to each of the three dosing arms of a single dosage of the IV bacteriophage therapy; the primary outcome of the Phase 1 arm of this study is treatment-related SAEs among only these 6 participants. In other words, safety data from only 6 subjects (2 in each dosing arm) was considered by the study investigators and the funding agency (NIH) as sufficient to proceed to Phases 2a/b. Assuming AchromoPhage is safe and tolerable in at least 90% of participants, a total sample size of 12 yields one-sided exact 90% confidence intervals with a width of no more than 31%. As some participants may unexpectedly drop out early, we will impose the restriction that randomization will continue until four participants in each treatment arm have completed all assessments planned for visits 1, 2, 3, and 4.

## Recruitment

Potential participants with cystic fibrosis CF will be identified through multiple avenues, including but not limited to:

1. Existing patient registries (e.g., CF clinic, research, and clinical databases)
2. Digital and print advertisements via social media platforms, CF societies, patient advocacy groups, and professional medical society listservs.
3. Referrals from healthcare providers at UPMC or other US health systems.
4. Self-referrals from patients expressing interest in participating.
5. Data from *Achromobacter* phage libraries (e.g., at the University of Pittsburgh or from other studies). All specimens used in this process will be deidentified, and no direct contact with participants will occur. Instead, the phage laboratories will communicate results to the original referring provider, who will determine whether to discuss the trial with the patient and make the appropriate referral to the study team.

There will be no cold-calling or direct outreach to potential participants. Where an individual is identified through a registry or database rather than by clinician or self-referral, the investigator will first consult the treating provider, who initiates the discussion with the patient; the study team contacts the patient only once the provider confirms their willingness to be approached. Participants are primarily identified in the outpatient setting, although hospitalized patients may also be eligible. To support enrollment from outside centers, prescreening may be completed remotely and travel and lodging costs are covered by the trial. Enrollment is expected to take approximately 24 months.

## Assignment of interventions: randomisation

### Sequence generation: who will generate the sequence

The allocation sequence was generated by the study statistician using Claude AI. The sequence is held within the University of Pittsburgh’s Research Electronic Data Capture (REDCap) system, and allocation is performed by the UPMC investigational pharmacy. To minimize bias and maintain allocation concealment, only pharmacy staff will have access to the randomization algorithm.

### Sequence generation: type of randomisation

An allocation sequence will be generated using permuted block randomization with random block sizes of 3 and 6, assigning participants to the three treatment arms in a 1:1:1 ratio. No stratification factors are used. Because randomization may extend beyond 12 participants, varying block sizes are used to reduce the predictability of the sequence. Details of the block structure are held in a separate document that is not available to personnel who enroll participants or assign interventions.

### Allocation concealment mechanism

Allocation is concealed through central randomization within REDCap. The randomization module is accessible only to investigational pharmacy staff; investigators and coordinators who identify, consent, and enroll participants have no access to the sequence and cannot view upcoming assignments. The allocation is released only after eligibility has been confirmed and the participant has been enrolled on Day 0, and is then communicated to the pharmacy for dose preparation.

### Implementation

Potential participants will be pre-screened and will then be enrolled into the study if the participant mees the rest of the eligibility criteria. Enrollment is carried out by the investigator or by study coordinators authorized by the institutional review board, and assignment to a treatment arm is carried out by the investigational pharmacy. The personnel who will enroll the study participants will not have access to the randomization sequence, which is accessible only to pharmacy staff*, and randomization will be conducted with the study specific REDCap system.

## Assignment of interventions: blinding

### Who will be blinded

This is an open-label trial. No blinding is used: participants, care providers, outcome assessors, and data analysts are all aware of the assigned route of administration. Allocation is concealed from all parties, including personnel enrolling participants until assignment, as described above, but is not concealed thereafter.

### How will be blinding be achieved

N/A – open label trial.

### Procedure for unblinding if needed

N/A – open label trial.

## Data collection and management

### Plans for assessment and collection of outcomes

Outcome data of interest (FEV1, CFQ-R, AEs, SAEs) will be collected at predetermined times by trained research personnel using IRB approved premade participant study diaries and case report forms (CRFs). Health-related quality of life is measured with the Cystic Fibrosis Questionnaire-Revised(CFQ-R), a validated instrument for persons with CF; the symptom diary is a study-specific instrument and has not been formally validated. Spirometry is performed according to American Thoracic Society and European Respiratory Society standards using the same device type throughout, and all data are transcribed from source documents into the electronic CRF.

### Plans to promote participant retention and complete follow-up

Study participants will be regularly contacted to ensure all study-related tasks are completed. Follow up safety evaluations will be conducted for the duration of the trial period of 42 days. Retention is supported by the option to complete the Day 21 and Day 42 visits remotely, home collection of respiratory specimens, and reimbursement of travel and lodging. Participants who discontinue the intervention are offered the opportunity to complete all remaining assessments, and those who withdraw are offered a single early exit visit, which may be conducted remotely or in part; if all follow-up is declined, medical records are reviewed for four weeks after the last dose to capture safety data.

### Data management

Study staff will store all data from the trial in the secure University of Pittsburgh REDCap system. Clinical specimens will be checked to ensure that all required and expected samples from each respective visit were collected. Participants will be assigned a randomly generated study ID. Linkage between participant and deidentified study ID will be stored in a University of Pittsburgh secured SharePoint cloud storage directory managed by the Center for Research on Health Care Data Center (CRHC-DC) at the University of Pittsburgh, which has obtained clearance for the maximum data classification level. Study records will be retained for at least seven years following the conclusion of the trial.

### Confidentiality

Personal health information (PHI) will be collected and then stored in a secure fashion within the electronic study database REDcap. All study information including CRFs, questionnaires, diaries, and patient specimens are kept strictly confidential. Records and specimens leaving the study site carry a coded identifier only, with the linkage held separately under restricted access. PHI is handled in accordance with HIPAA and is not disclosed outside the study team except to regulatory and oversight bodies bound by confidentiality obligations, or as required by law.

## Statistical methods

### Statistical methods for primary and secondary outcomes

We will perform statistical modeling, but formal inference will be limited, and no formal between-group comparisons will be performed for primary or secondary outcomes. We will focus on estimation and precision in outcome assessments, as is appropriate for early stage trials (45). Confidence intervals (CIs) will be used to gauge the precision of estimates. For binary outcomes (e.g. safety/tolerability), we will construct a random intercept (RI) logistic regression model with main effects for treatment modality (IV, inhaled, and combination), week as a categorical predictor, and the interaction between treatment modality and week. For continuous outcomes (e.g., changes in bacterial abundance or FEV1; transformations applied as needed), RI linear models with the same fixed effects will be fitted. Both logistic and linear RI models will include a random effect for participant to control for repeated measures.

For the primary safety outcome, back-transformed estimates of incidence at each time point along with one-sided 90% CIs, for all modalities combined as well as separately for each modality, will be obtained via least-squares means (LS-means) estimation from the interaction-based RI logistic model. LS-means estimation will similarly be employed for secondary, continuous outcomes from the interaction-based RI linear models, and all estimates will be accompanied by two-sided 90% CIs. However, formal between-arm inference will still be limited. Comparisons of outcomes between modalities over time will not employ formal hypothesis testing. Rather, comparisons between arms at each time point will be constructed via difference in LS-means estimation and represented by pairwise, 2-sided 90% CIs for the difference in incidence (or means for secondary outcomes) between modalities, with no adjustment made to the confidence level for multiple comparisons given the pilot nature of the study. In the case that random effects models are unstable or do not converge in light the small number of participants (N=12), we will summarize results at each time point for each treatment modality using descriptive statistics. For binary outcomes (e.g., safety/tolerability), estimates of incidence will be reported as unadjusted proportions with exact 1-sided 90% confidence intervals, stratified by treatment arm and time.

Assuming phages are safe and tolerable in ≥ 90% of participants and given a combined sample size of N=12 participants, 1-sided exact 90% CIs for these binary measures will have CI widths of at most 31% (90% CI: 69%-100%). For continuous endpoints, we expect to observe 2-sided 90% CI widths of at most 1.04 standard deviations, assuming a moderate within-participant correlation of ρ=0.5. These levels of precision are reasonable considering our sample size, and we expect even greater precision because of multilevel/repeated measures modeling. Outcome estimates will inform the design of future trials.

For continuous secondary outcomes, point estimates of change from baseline will be reported descriptively using unadjusted means and standard deviations and accompanied by 2-sided 90% confidence intervals, stratified by treatment arm and follow-up time point. CFQ-R quality of life domain scores will be computed and standardized. Separately for each domain, linear mixed models will be fit as a function of week (categorical), study arm, and week-by-arm interaction; a random intercept for subject will be included. We will use LS-means estimation to calculate point estimates and corresponding two-sided 90% CIs for the mean domain score at each time point. Using these interaction models, we will derive estimates for arm-specific longitudinal trajectories, as well as the trajectory for the sample as a whole. The data from the biological endpoints (e.g., phage PK, abundance, susceptibility profiles, neutralizing antibodies) will be considered descriptive. However, we will assess differences in phage and bacterial abundance between treatment arms using unpaired two-tailed t-tests. We will compare phage and bacterial abundance between participants using analysis of variance (ANOVA) and will assess changes in these variables over time in each participant using linear regression. We will use t-test and ANOVA to assess differences in serum neutralization both within and between participants.

### Who will be included in each analysis

Analysis will be performed on the intention-to-treat principle, with participants being analyzed in each respective study arm they were originally assigned to.

### How missing data will be handled in the analysis

We will make every attempt to document all reasons for missing data. Reasons for intermittently missing data will be investigated. The proposed likelihood-based statistical procedures (RI models) allow for analysis of every participant’s data, regardless of attrition, and provide valid results when data are “missing at random” (MAR). If the MAR assumption is found to be questionable, we will fit shared parameter models that allow for non-ignorable missingness.

### Methods for additional analyses (e.g. subgroup analyses)

Subgroup analyses are not planned. An exploratory analysis of the primary endpoint will summarize event rates within arms across time points, stratified by CTCAE category and grade, for categories and grades in which at least one event occurs, in order to identify which event types drive the composite endpoint.

### Interim analyses

There is no planned interim analysis of both the primary and secondary endpoints.

### Protocol and statistical analysis plan

The statistical analysis is described in full within the protocol

## Oversight and monitoring

### Composition of the coordinating centre and trial steering committee

The University of Pittsburgh serves as the data coordinating center, and the Sponsor-Investigator holds overall responsibility for trial conduct and safety oversight. A study oversight team of the Sponsor-Investigator, co-investigators, site investigators, and key study personnel meets weekly to review enrollment, retention, adverse events, unanticipated problems, and protocol deviations. There is no separate trial steering committee or endpoint adjudication committee; adverse events and pulmonary exacerbations are adjudicated by the investigator in consultation with the Sponsor-Investigator.

### Composition of the data monitoring committee, its role and reporting structure

This trial is monitored by a DSMB convened at the University of Pittsburgh, independent of the study investigators, whose members attest in writing to the absence of conflicts of interest. Membership may include experts in infectious diseases, pulmonary medicine, and clinical trials or biostatistics, and includes two members external to the University of Pittsburgh and UPMC. The DSMB meets before trial initiation, approximately four times during the trial, and once at its conclusion, reviewing safety updates, adverse events, unanticipated problems, and study progress, and makes recommendations on continuation, modification, or termination. After each meeting the Sponsor-Investigator receives a formal letter from the chair, which is shared with the IRB and the Cystic Fibrosis Foundation. Further detail is provided in the DSMB charter.

### Frequency and plans for auditing trial conduct

Independent monitoring is provided under contract by the Education and Compliance office for Human Subject Research at the University of Pittsburgh. Monitors conduct periodic site visits at which they review consent forms, electronic case report forms, source data, specimen and study product records, and site regulatory files. The site provides direct access to all trial-related facilities, source documents, and reports for monitoring and auditing by the sponsor and inspection by regulatory authorities, including the FDA. The IRB additionally conducts continuing review.

### Protocol amendments

Any protocol amendments will be submitted for review to each participating IRB for approval prior to implementation, and substantive amendments are also submitted to the FDA under the IND and reflected in the ClinicalTrials.gov record. If an amendment affects participation or the consent form, participants will be notified and asked to sign updated consent forms.

### Dissemination policy

Results of the trial will be submitted to clinicaltrials.gov within 1 year of study completion, and the study staff plan to submit the results of the trial for publication in a peer-reviewed scientific journal. Study results may be discussed/presented at relevant conferences. Participant PHI will be de-identified in all data released from the trial.

### Ancillary and post-trial care

Study participants are monitored for adverse events from the time of informed consent through Visit 7, four weeks after the final phage dose. We will not provide routine ancillary or post-trial care. However, if any complications are thought to be caused by the trial, the participants will be followed until the event resolves or stabilizes, with longer-term follow-up coordinated through routine clinical care where appropriate. Because AchromoPhage is investigational and not approved for clinical use, no continued access to the product is provided after the trial ends.

## Discussion

The AchromoPhage Trial is a first-in-human evaluation of a bacteriophage cocktail developed specifically for *Achromobacter*, a poorly studied CF pathogen associated with accelerated lung function decline and poor outcomes. Phage trials in cystic fibrosis to date have targeted more common pathogens, principally *P. aeruginosa*, leaving *Achromobacter* — for which antibiotic options are frequently exhausted — without a comparable development pathway. Despite renewed enthusiasm in phage therapy, the field has not been matched by the level of scientific rigor expected of therapeutic development. Instead of large prospective trials preceding clinical use, the field has expanded through compassionate use and case series, with systematically evaluation of safety and efficacy beginning only recently (17,20,34,56). Because existing safety data therefore derive largely from uncontrolled, last-resort administrations, we designed this trial around safety and tolerability, with secondary and exploratory outcomes — lung function, quality of life, pharmacokinetics and pharmacodynamics, and the humoral response — intended to inform the dose, route, and endpoints of later trials powered for efficacy. The trial also demonstrates that academic investigators can carry a novel antimicrobial from phage isolation through IND sponsorship to clinical execution, a pathway that may matter increasingly for pathogens that attract little commercial interest.

Safety monitoring for phage therapy in pwCF is particularly challenging, as the events of interest (cough, increased sputum production, fever etc.) are also features of the underlying lung disease. We address this by soliciting a prespecified list of symptoms at every study contact, collecting symptom diaries and home temperature measurements around dosing visits, and tracking expected disease-related events separately from adverse events, with attribution adjudicated by the investigator against each participant’s own baseline. The severity of the underlying lung disease poses a further dilemma: individuals with advanced lung disease may stand to benefit most from an investigational intervention yet are more likely to be excluded from early trials due to the severity of their underlying illness. In this first-in-human trial, and in discussion with the FDA, we elected to exclude participants with an FEV1 below 40% of predicted; evaluating AchromoPhage in this group is a priority once the safety of the cocktail is established.

Beyond safety data, little data is known about the optimal dose, route, and duration of phage therapy, and animal and in vitro pharmacokinetic models remain sparse while large-scale human studies do not exist. Dosing is further complicated by the self-replicating nature of phage, which amplifies at the site of infection in the presence of susceptible bacteria, so the administered dose may not reflect the exposure ultimately achieved. We chose our dosing schedule (one dose a week for three weeks) partly because it gives adequate time between doses and allows for monitoring of AEs in a prolonged timeframe; more frequent intravenous dosing would also require indwelling catheterization and home health support. The type of nebulizer used for phage delivery into the lungs is also important: studies have shown that phage delivery device may affect the viability of phage after nebulization(44,57), and we selected a vibrating mesh nebulizer on this basis. These parameters may not apply to different phage products, as phage characteristics such as receptor bacterial surface proteins, replication time, and burst size vary widely across phages.

The design addresses several of these gaps directly. Randomizing participants among inhaled, intravenous, and combined administration allows the three routes to be compared within a single protocol, and serial specimens collected before and after each dose will support estimation of phage concentrations in blood and the airway, bacterial burden in sputum, and the emergence of phage resistance. Questions this trial cannot resolve — optimal duration, the role of the innate and adaptive immune response to phage, and the best route for other infection types — will require subsequent studies.

These considerations argue for studying phage in the populations who will ultimately receive them. Unlike conventional antimicrobials, phage pharmacokinetics are driven by pharmacodynamics: in the absence of a susceptible bacterial host, phage are cleared without amplifying, so exposure measured in a healthy volunteer bears little relation to exposure at an infected site. Healthy volunteer studies can therefore contribute data on tolerability and immunogenicity, but little of what most needs to be established about dose, route, and duration. Given how rapidly resistance is outstripping the treatment options available to pwCF, we elected to conduct this first-in-human evaluation in participants with chronic *Achromobacter* infection, using dose escalation, prespecified stopping rules, and independent safety monitoring to protect them. This approach allows safety data to be generated alongside the pharmacokinetic, pharmacodynamic, and microbiologic data needed to design the next trial, in a population for whom phage therapy, if it works, would matter most.

## Data Availability

All data produced in the present study are available upon reasonable request to the authors

## Trial status

This is Protocol Version 2.0, dated August 26, 2026. Recruitment for the study began on August 1, 2026 and is ongoing. We expect recruitment and follow-up to be completed by 2028.

## Abbreviations

AE: Adverse events
ALT: Alanine Aminotransferase
ANOVA: Analysis of Variance
AST: Aspartate Aminotransferase
CBCd: Complete Blood Count with differential
CF: Cystic Fibrosis
CFQ-R: Cystic Fibrosis Questionnaire-Revised
CI: Confidence Interval
CRF: Case Report Form
CRHC-DC: Center for Research on Health Care Data Center
DRE: Disease Related Event
DSMB: Data and Safety Monitoring Board
CMP: Complete Metabolic Panel
DSA: Donor Specific Antibodies (also known as HLA class I and II Antibody Screen)
eCRF: Electronic Case Report Form
FDA: Food and Drug Association
FEV1: Forced Expiratory Volume in 1 second
IND: Investigational New Drug
IRB: Institutional Review Board
IV: Intravenous
LS-means: Least Square means
MAR: Missing at Random
MDRO: Multi-drug resistant organism
P3: Pittsburgh Phage Program
PBS: Phosphate Buffered Solution
PFU: Plaque forming unit
PHI: Protected Health Information
PRA: Panel Reactive Antibodies
pwCF: People with cystic fibrosis
REDCap: Research Electronic Data Capture
RI: Random Intercept
SAE: Serious Adverse Events
SOT: Solid organ transplant
ULN: Upper Limit of Normal
UPMC: University of Pittsburgh Medical Center
V/Q: Ventilation/Perfusion

## Acknowledgements

Not applicable

## Declarations

### Authors’ contributions

Authorship was given to individuals who made a substantial contrinution to study design, study conception, data analysis/interpretation; drafting or revising the manuscript; and agreed to accountability for the manuscript.

GH is the Sponsor Investigator; he co-conceived the study, co-developed the proposal, and led protocol development.

DVT is co-Principal Investigator; she co-conceived the study, co-developed the proposal, identified and manufactured the phages; her lab will perform the proposed assays.

MS is sub-investigator; she contributed to the conception/design of the study and substantively revised the manuscript. She will also assist in interpreting the data.

RS is sub-investigator; he co-conceived pharmacokinetic parameters and timepoints for the trial. He substantively revised the manuscript. He will also assist in interpreting the data.

AZ is sub-investigator, she contributed to the conception/design of this study and substantively revised this manuscript

KR is sub-investigator; she contributed to the conception/design of this study and substantively revised this manuscript.

JP is sub-investigator; he contributed to the conception/design of this study and substantively revised this manuscript.

FS is sub-investigator; she contributed to the conception/design of this study and substantively revised this manuscript

SA is sub-investigator; she contributed to the conception/design of this study and substantively revised this manuscript. She is the site leader at UCSD.

SR is sub-investigator; he conceived of the biostatistics, contributed to the conception/design of this study, and substantively revised this manuscript.

SSB is clinical research coordinator; she coordinated the clinical research. She substantively revised this manuscript.

BB is program manager or the Pittsburgh Phage Program. She coordinated the clinical research. She substantively revised this manuscript.

KHK is sub-investigator and regulatory director; she contributed to the conception/design of the study and substantively revised the manuscript. She will also assist in interpreting the data.

JM will assist in gathering and interpreting data and wrote the protocol paper.

All authors read and approved the final manuscript.

### Availability of data and materials

Upon completion of the study, final data may be supplied on request, following submission of a plan for data use, approval of the plan by the NIH, and execution of required institutional agreements.

### Ethics approval and consent to participate

This study utilizes a centralized IRB. The study protocol and centralized IRB conclusion will be reviewed and adopted by each site’s relevant local IRB prior to participation. All volunteers require written informed consent to participate

